# Conflicts of interest and physicians’ attitudes towards hydroxychloroquine as a treatment against COVID-19 A replication and extension of Roussel & Raoult (2020)

**DOI:** 10.1101/2021.10.18.21265135

**Authors:** Louis Freget, Matthieu Mulot, Michael Rochoy, Valentin Ruggeri, Céline Schöpfer, Florian Cova

## Abstract

Hydroxychloroquine (HCQ) and its use as a treatment against COVID-19 have been at the center of heated debates. Some claim that physicians’ hostility towards HCQ was partly orchestrated by rival pharmaceutical companies seeking to promote their own treatment. In favor of this hypothesis, Roussel and Raoult (2020) have presented the results of a study in which they find a perfect positive correlation (ρ = 1.00) between French physicians’ attitudes towards HCQ and their conflict of interest with Gilead Sciences - the company that has promoted Remdesivir (REM) as a treatment against COVID-19. However, Roussel and Raoult’s study suffers from serious methodological shortcomings, among which is the fact that the statistical methods they employed might tend to artificially inflate correlations. In this study, we use a similar method and sample, but correct for their study’s original shortcomings: we provide a detailed, pre-registered method for collecting and coding data, computer inter-rater agreement and use a wide array of appropriate statistical methods to achieve a more reliable estimate the association between conflicts of interest and physicians’ attitudes towards HCQ. We conclude that Roussel and Raoult’s conclusion was misguided and that financial conflicts of interest were not the main predictors of the attitudes of physicians when compared to other factors, such as academic affiliation. Moreover, compared to other pharmaceutical companies, there was no specific link between attitudes towards HCQ and conflicts of interest with Gilead Sciences.

## Introduction

Against the backdrop of the COVID-19 pandemic, numerous treatments have been put to test. Among those, one has stirred many debates: hydroxychloroquine (hereafter: HCQ). In France, this drug was notably promoted by Didier Raoult, a world renowned microbiologist, and his team, on the basis of *in vitro* and observational studies. However, their proposal failed to convince other scientists, which led to a scientific controversy that, far from being limited to the scientific field, engulfed French society as a whole [1,2,3]. Some could argue that scientists’ opposition to HCQ was driven by the lack of robust scientific evidence in its favor [4,5,6]. However, others have claimed that its demise was the product of a smear campaign (a “war against chloroquine”) organized by pharmaceutical enterprises [7]. More specifically, it has been claimed that HCQ was attacked to promote another treatment: Remdesivir (hereafter: REM). This makes the producer of REM, Gilead Sciences, a prime suspect in HCQ’s fall from grace [8].

In support of this narrative, Roussel and Raoult [9] set out to investigate “the links of interest between Gilead Sciences, producer of Remdesivir, and the doctors who took a stand for or against hydroxychloroquine.” [7] Starting from a list of members of the French Council of Teachers in Infectious and Tropical Diseases (CMIT), they identified 98 medical researchers and, for each of them, (i) identified their conflict of interests with Gilead Sciences and pharmaceutical enterprises in general, and (ii) evaluated their attitude towards HCQ. Their analysis returned a *perfect* inverse correlation between the two variables: ρ = −1.00, *p* = 0.02, *N* = 5, suggesting that conflict of interests with Gilead Sciences played a major role in physicians’ attitudes towards HCQ.

However, Roussel and Raoult’s study suffers from major statistical and methodological shortcomings that have already been described in detail [10]. On the statistical side, Roussel and Raoult chose to aggregate their 44 data points into 5 data points, leading to a massive loss of information that might artificially inflate their correlations. This problem could be solved by reanalyzing Roussel and Raoult’s data, but our requests for the data remained unanswered.

Even if we had access to these data, there would still remain serious methodological issues in Roussel and Raoult’s study. First, their method for coding is underspecified: we don’t know what they did when the two coders came to different verdicts, or when the same physician expressed different attitudes in different circumstances. Moreover, they do not provide any statistical information about inter-rater agreement. Second, the categories they used to code physicians’ attitudes towards HCQ were problematic. Indeed, the categories were ambiguous about the kind of attitudes they were trying to measure: in certain cases, the categories seem to assess whether physicians believe in the effectiveness of HCQ against COVID-19 (“having recognized a positive effect of hydroxychloroquine”, “expressing the need for more studies for making any comment of the efficiency of the treatment”) and whether they promote its use in fighting COVID-19 (“having expressed a call for generalization of the use of hydroxychloroquine”). At other times, they assess physicians’ attitudes towards the mediatization of the debate about HCQ (“expression of anger towards the mediatization of hydroxychloroquine”). Finally, in certain cases, the criterion can be hopelessly vague (“expressing negative comments about hydroxychloroquine”): does informing the public about the potential negative side-effects of HCQ count as “negative comments”? Thus, Roussel and Raoult’s coding categories mix several dimensions and criteria of assessments: being against the mediatization of the debate over HCQ is not the same as claiming that HCQ is ineffective. And making “negative comments” about HCQ (for example, by pointing out its potential negative side-effects) is not the same as being against the mediatization of the scientific debate, or being against its use in treating COVID-19. Additionally, the categories used by Roussel and Raoult are not symmetrical: while the definition of negative attitudes points towards physicians’ emotional states (“expressing anger”), there is no such thing for the definition of positive attitudes. On the contrary, positive attitudes are more likely defined in terms of scientific data (“reporting a successful use of the treatment in the physician’s facility”) than negative attitudes.

Thus, Roussel and Raoult’s methodology sheds doubt on their conclusion, according to which French physicians’ conflicts of interest with Gilead Sciences could explain their opposition to HCQ. As this possibility raises serious worries about scientific integrity, our aim was to replicate Roussel and Raoult’s study while improving on the shortcomings and limitations we just detailed. Among such improvements is the fact that all materials, data and analysis script for the present study are available at osf.io/uahq6/

## Materials and methods

Following Roussel and Raoult’s procedure, we started from a list of 115 CMIT members. For each physician on the list, we collected (i) their conflicts of interest with Gilead Sciences, Sanofi and pharmaceutical enterprises in general, and (ii) their attitudes towards HCQ and REM as treatment against COVID-19. Data about conflicts of interest were collected through the [Transparence Santé](https://www.transparence.sante.gouv.fr/) public database. Data about attitudes were collected by having researchers search for the physician’s name through Google News and Google Scholar. Search was limited to two time periods:

- Period 1: from February 25^th^, 2020 (the day Didier Raoult published his video in which he claimed that COVID-19 could be cured using HCQ) to May 5^th^, 2020 (the day Roussel and Raoult submitted their original paper).
- Period 2: from August 26^th^, 2020 (the day Fiolet et al.’s meta-analysis on the effect of HCQ was published online) to December 31^st^, 2020 (when we decided to start this project).

Once the data were collected, each coder was asked to code the interventions they themselves collected. Then, the coding of the interventions was assigned to another coder who coded the same interventions again. A third coder settled the disagreements.

The coding categories were the following:

2 = ‘Very favourable’, defined as ‘having expressed a call for generalization of the use of hydroxychloroquine/remdesivir, reporting a successful use of the treatment in the physician’s facility, or claiming that the treatment works against COVID-19’.
1 = ‘Favourable’, defined as ‘having expressed positive attitudes (i.e. hopes, probability of efficiency) about the use of hydroxychloroquine/remdesivir, while waiting for more results for taking further position’.
0 = ‘Neutral’, defined as ‘expressing the need for more studies for making any comment on the efficiency of the treatment’.
-1 = ‘Unfavourable’ was defined as ‘having expressed negative attitudes (i.e. suspicion, probability of inefficiency) about the use of hydroxychloroquine/remdesivir, while waiting for more results for taking further position’.
-2 = ‘Very unfavourable’ was defined as ‘having expressed a call for interdiction of the use of hydroxychloroquine/remdesivir, or reporting an unsuccessful use of the treatment in the physician’s facility, or claiming that the treatment does not work against COVID-19’.

Our coding scheme differs from the one used by Roussel & Raoult (2020) in several respects. First and foremost, we tried to make our coding categories symmetrical (‘Very unfavourable’ mirrors ‘Very favourable’, while ‘Unfavourable’ mirrors ‘Favourable’). Second, we tried to get rid of emotional terms (such as ‘expressing anger’) that might have made the coding procedure more subjective. And finally, but most importantly, we tried to be more specific about the standard according to which an opinion about the use of HCQ and REM should be considered ‘positive’ or ‘negative’: their effectiveness in treating and/or preventing COVID-19.

## Results

### Inter-rater agreement

A total of 252 interventions were collected and coded. Coders agreed in 67% of cases (unweighted Cohen’s kappa for categorical data: 0.57 [0.50, 0.65]). Leaving aside interventions for which one coder rated NA, we found a Pearson’s correlation of *r* = 0.83 [0.79, 0.87] between coder 1 and coder 2’s ratings. Overall, inter-rater agreement was satisfying. See the Appendix for details.

### Physicians’ attitudes

From coders’ ratings, we computed each physician’s attitudes towards each treatment (HCQ or REM) for each time period (1 or 2). When a same physician made several interventions and some of them received different ratings, we computed the average rating and then (i) selected the most extreme negative rating if the average was below 0, (ii) selected the most extreme positive rating if the average was above 0, (iii) settled for a score 0 of the average was 0. We chose this computation method to obtain a final score that would be discrete (−2, −1, 0, 1 and 2) and would mirror Roussel & Raoult’s own scoring system. See the Appendix for an analysis of average attitude scores.

Table 1 shows the total number of physicians for which we were able to compute an attitude score for each treatment and each period, along with mean and standard deviations for attitude scores. Compared to the first period, approbation of the use of both molecules dropped in the second period: physician’s most extreme attitudes were significantly lower in the second period compared to the first for both HCQ (*t*(68) = 3.68, *p* < .001) and REM (*t*(23) = 2.14, *p* = 0.043). This suggests that physicians’ attitudes were not set in stone, but sensitive to the scientific evidence gathered between the two periods.

**Table 1.** Number of physicians expressing an attitude towards HCQ and REM in the first and second period, along with mean and standard deviations for attitude scores.

|  | 1st period | 2nd period |
| --- | --- | --- |
| HCQ | $N = 50$<br>$\mu = 0.32$, $SD = 1.15$ | $N = 20$<br>$\mu = -0.95$, $SD = 1.64$ |
| REM | $N = 13$<br>$\mu = 0.46$, $SD = 0.78$ | $N = 12$<br>$\mu = -0.58$, $SD = 1.56$ |

Only for physicians’ attitudes towards HCQ in the first period did we reach a reasonable sample size (*N* = 50). We thus focused our analysis on these data, leaving aside physicians’ attitudes towards HCQ in the second period, and physicians’ attitudes towards REM during the first and second period. Note however that physicians’ most extreme attitudes towards REM were not significantly correlated with the amount of money they received from Gilead (Pearson, first period:, *r*(13) = .02, *p* = .95; second period: *r*(12) = -.00, *p* = .99).

### Conflict of interests as predictors of physicians’ attitudes towards HCQ

To assess the relationship between physicians’ conflict of interests with Gilead Sciences and their attitude towards HCQ, we first used the same statistical approach as Roussel and Raoult. We computed the average amount of money received from Gilead for each possible attitude towards HCQ (see Table 2). Then we computed a Spearman rank-correlation between attitudes treated as a rank and the average amount of money received by physicians, for a total of 5 data points. The correlation is extremely strong (*ρ* = −0.90 ; R^2^ =0.67). However,this method tends to overlook variability within each category of attitudes, thus contributing to artificially inflating correlations.

**Table 2.** Average, median, minimum and maximum amount of money received from Gilead in function of physicians’ attitudes towards HCQ (first period).

| Attitude | N | Mean | Median | Minimum | Maximum |
| --- | --- | --- | --- | --- | --- |
| Very unfavourable | 5 | 14293 | 12335 | 122 | 30365 |
| Unfavourable | 4 | 14182 | 2545 | 26 | 51612 |
| Neutral | 19 | 4730 | 1945 | 0 | 31731 |
| Favourable | 14 | 10486 | 800 | 0 | 52012 |
| Very favourable | 8 | 47 | 0 | 0 | 375 |
| Did not take position | 65 | 4980 | 1687 | 0 | 48086 |

As recommended by Piepho [10], we rather used individual physicians as data points. Figure 1 shows that this gives a more nuanced image of the relationship between physicians’ attitudes and conflicts of interest. Using physicians’ attitude towards HCQ as a dependent variable and the total amount of money they received from Gilead, the total amount of money they received from Sanofi, the total amount of money they received from pharmaceutical companies, the percentage of total conflict of interests coming from Gilead and the percentage of total conflicts of interest coming from Sanofi as separate predictors, we assessed the strength of this relationship using two different methods: Pearson correlation, and regression analysis (OLS). Results are presented in Table 3.

**Table 3.** Physicians’ attitudes towards HCQ (first period) in function of various measures of conflict of interests (two methods: Pearson correlation, OLS). For the %Gilead and %Sanofi column, physicians who received no money from pharmaceutical industries were excluded, leaving us with *N* = 49 physicians.

|  | *Gilead* | *Sanofi* | *Total* | *% Gilead* | *% Sanofi* |
| --- | --- | --- | --- | --- | --- |
| **Pearson** | $r = -0.24$<br>$p = 0.098$ | $r = 0.00$<br>$p = 0.98$ | $r = -0.36$<br>$p = 0.011$ | $r = -0.12$<br>$p = 0.39$ | $r = 0.24$<br>$p = 0.092$ |
| **Regression**<br>**(OLS)** | $R^2 = 0.06$ | $R^2 = 0.00$ | $R^2 = 0.13$ | $R^2 = 0.02$ | $R^2 = 0.06$ |

**Figure 1.**
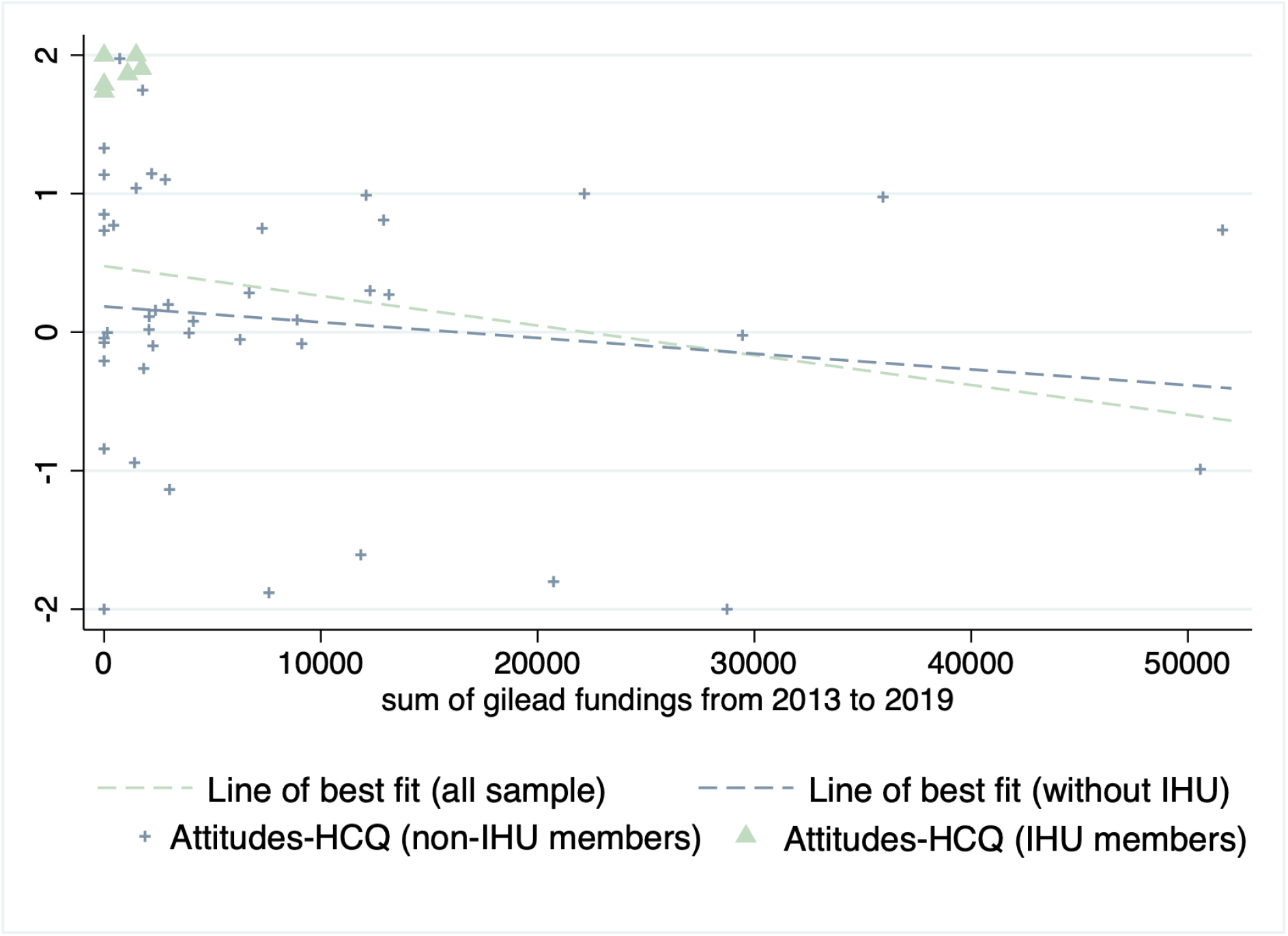
Plotting the relationship between most extreme attitudes towards HCQ and gilead fundings among IHU members and non IHU members. Data was jitterized in order to circumvent overplotting. The same plot with average attitudes is displayed in the appendix (Figure S1).

The correlation between participants’ attitudes and their conflict of interests with Gilead Sciences was small and non-significant. There was however a significant relationship between physicians’ attitudes and their total conflicts of interest. Neither was there a significant association between attitudes and the ratio of gilead fundings to total fundings. Thus, there does not seem to be any privileged link between Gilead and attitudes towards HCQ, compared to pharmaceutical companies in general.

A look at Figure 1 suggested that both correlations were partly driven by a small cluster of physicians with very low conflicts of interest and very positive attitude towards HCQ. It turned out that most of these physicians were members of the IHU Méditerannée Infection (IHU-MI) and that being a member of the IHU-MI was a very strong predictor of attitudes towards HCQ (β = 0.54, *p* < 0.001, *R*^*2*^ = 0.30). Once members of the IHU-MI were excluded from analysis, correlations between physicians’ attitudes and their conflicts of interests became weaker and non-significant when considering gilead fundings (*r*(42) = −0.15, *p* = 0.345), and non or marginally significant when considering total fundings (*r*(42) = −0.29, *p* = 0.054).

## Discussion

Overall, our results suggest no robust relationship between physicians’ conflicts of interest with Gilead Sciences and their attitudes towards HCQ (see the Appendix for a series of sensitivity and multivariate analyses). In fact, the share of variance explained in physicians’ attitudes by their conflicts of interest with Gilead Science (6%) was greatly inferior to the share of variance explained by other factors, such as physicians’ affiliation to the IHU-MI (30%). This suggests that, while trying to explain physician’s attitudes, we should not only look at *financial* conflict of interest, but also at *institutional* factors. As a researcher [11] once wrote: “*it is naive to consider that conflict of interests are limited to those that come from fundings: there are many others. Among the most important are ideological conflicts. Some people take a religious stance on scientific theories - an approach that often allowed them to develop their thought and progress in their career. Questioning these theories threaten their beliefs and can sometimes trigger incredibly violent reactions. (*…*) There is of course another source of influence: affective variables. We look more kindly upon a theory (even one that runs against our own worldview) when it comes from someone we love or respect)*.”

## Data Availability

All data produced are available online at: osf.io/uahq6/

## Author contributions

Louis Fréget: Conceptualization, Methodology, Data Collection, Data Analysis, Writing - Original Draft

Matthieu Mulot: Conceptualization, Data Collection, Writing - Review & Editing

Michaël Rochoy: Conceptualization, Data Collection, Writing - Review & Editing

Valentin Ruggeri: Conceptualization, Data Collection, Writing - Review & Editing

Céline Schöpfer: Data Collection, Writing - Review & Editing

Florian Cova: Conceptualization, Methodology, Data Collection, Data Analysis, Writing - Original Draft

## ONLINE APPENDIX

The present appendix is structured as follows.

In Appendix 1, we detailed the data collection and coding procedures.

In Appendix 2, we led multivariate and mediation analyses as specified in the pre-registration plan. Neither the H index nor IHU affiliation are suitable mediators for the relationship between fundings and attitudes. However, when controlling by these variables, the association between gilead funding and attitudes towards HCQ disappears and that between opinion and total fundings becomes only marginally significant.

Figure S1 is a similar figure as Figure 1 but with average attitudes instead of the most extreme attitudes.

The rest of this appendix is dedicated to five ranges of sensitivity analyses which we used to guarantee the robustness of our results.

In Appendix 3, we assessed the relationship between the various measures of fundings and physicians’ attitudes towards HCQ using an ordered probit regression which might be more suited for our discrete outcome. We find the same results as in our main analysis. While the association between total fundings and attitudes is significant in bivariate models, none of the other components of the fundings significantly correlates with attitudes. This holds even when excluding the members of the IHU and controlling by the h-index.

In Appendix 4, as specified in the pre-registration, we ran the correlational analyses of Section 4 a second time with physicians’ attitudes computed as their average attitude across multiple interventions (rather than selecting the most extreme attitude). While using this approach makes the association between Gilead fundings and attitudes towards HCQ become significant, there is still a stronger association between total fundings and attitudes towards HCQ, and the association between the ratio of Gilead fundings and attitudes toward HCQ is still non-significant.

In Appendix 5, in an effort to increase the sample size, we created three dummies, one for having expressed a positive opinion, a second for expressing a neutral opinion, and a third one for expressing a negative opinion towards HCQ. We then looked at the association between each measure of fundings and these indicators using a logit model. The results of this exercise are in line with our core findings. There is an association between Gilead fundings and expressing a negative attitude towards HCQ but (i) there is still a much stronger association between total fundings and attitudes, (ii) the association between the ratio of gilead fundings to total fundings and expressing a negative opinion towards HCQ is still non-significant.

In Appendix 6, we used a mixed (multi-level) model to account for the hierarchical structure of the data. When nesting the data at the level of the IHU, the association between Gilead fundings and average opinions towards HCQ became marginally significant. The association between gilead fundings and the most extreme attitudes was still non-significant.

In Appendix 7, we checked whether h-index and Gilead fundings could be suitable mediators of the significant relationship we found in Annex 1 between Gilead fundings and average attitudes towards HCQ. It is not the case.

In Appendix 8, we included two other variables in our dataset : age (which was simply estimated by computing 2021 - their year of birth), and whether the physician is a professor or a mere medical doctor. These are other proxies of the competence/prestige of the physicians. Controlling by these variables does not change our main result and they are not suitable mediators.

## Appendix 1. details of the data collection and coding procedure

### 3.1 List of physicians

A list of the 115 members of the CMIT (Collège des Universitaires de Maladie Infectieuse et Tropicale) was kindly provided by the CMIT itself.

### 3.2. Data collection for physicians’ interventions

The data collection task for physician’s interventions was distributed among a total of 6 coders (CS, FC, LF, MM, MR and VR). Each physician on the CMIT list was randomly assigned to one coder (a few were accidentally attributed to two coders). Coder FC was the one in charge of assigning physicians to coders and compiling the works of other coders.

For each physician assigned to them, each coder were asked to go through Google News and Google Scholar to collect public interventions (e.g. interviews, petitions) or scientific articles in which the physician expressed an opinion or formulated a judgment about the efficiency of Hydroxychloroquine (HCQ)^1^ or Remdesivir (REM) in treating and/or preventing COVID-19.

Periods of interest for data collection were:

### 3.3. Coding - Phase 1

Once the data were collected, each coder was asked to code the intervention they themselves collected. The coding categories were the following:

2 = ‘Very favourable’, defined as ‘having expressed a call for generalization of the use of hydroxychloroquine/remdesivir, reporting a successful use of the treatment in the physician’s facility, or claiming that the treatment works against COVID-19’.
1 = ‘Favourable’, defined as ‘having expressed positive attitudes (i.e. hopes, probability of efficiency) about the use of hydroxychloroquine/remdesivir, while waiting for more results for taking further position’.
0 = ‘Neutral’, defined as ‘expressing the need for more studies for making any comment on the efficiency of the treatment’.
-1 = ‘Unfavourable’ was defined as ‘having expressed negative attitudes (i.e. suspicion, probability of inefficiency) about the use of hydroxychloroquine/remdesivir, while waiting for more results for taking further position’.
-2 = ‘Very unfavourable’ was defined as ‘having expressed a call for interdiction of the use of hydroxychloroquine/remdesivir, or reporting an unsuccessful use of the treatment in the physician’s facility, or claiming that the treatment works against COVID-19’.^2^

This has several implications. The first is that pointing out the possible harmful side-effects of HCQ did not count as a negative opinion, as long as the physician did not also express serious doubts about the positive main effects of HCQ on treating COVID-19. This allowed us not to run into conflicts whenever we found a physician claiming both that HCQ was effective against COVID-19 and that HCQ could potentially have dangerous cardiac side-effects. The second is that physicians who criticized the methodology of studies supporting the use of HCQ (or REM) or deplored the mediatic frenzy about HCQ while still acknowledging the possibility that HCQ might be effective were rated as ‘neutral’ rather than ‘unfavourable’. Indeed, criticizing a study, or telling people to be more cautious is not the same as doubting or denying the effectiveness of a molecule. As mentioned in the introduction, Roussel and Raoult (2020)’s method did not allow them to distinguish these different dimensions.

Coding was not blind, in the sense that it was impossible to hide the identity of physicians to coders: physicians were clearly identified in the material to be coded (such as interviews). However, all coders but LF were absolutely ignorant of the amount of benefits each coder had received from pharmaceutical industries. LF was the exception because he handled the data collection for benefits. However, to the extent that data collection was automated, it can be considered that LF did not have full knowledge of the physicians’ conflicts of interest. Moreover, none of the authors are infectiologists and most of the authors are not physicians.

### 3.4. Coding - Phase 2

In a second phase, each physician was assigned to a second (and sometimes to a third) coder. The second coder had access to all interventions collected by the first coder, but not to their coding. The second coder had to rate all interventions and had the possibility to add new interventions if they found some that had eluded the first coder. When new interventions were added, they were also sent to the first coder to collect their coding.

In addition to the five coding categories used in the first phase, a new category was included : ‘NA’. ‘NA’ was to be used by the second coder whenever they felt like the intervention collected by the first coder was not precise enough to infer the physician’s opinion.

### 3.5. Coding - Phase 3

Once first and second coders had finished their task, FC and LF compiled all their answers in a single document. Whenever there was a disagreement between the first and the second coders, FC and LF went back to the physician’s intervention and settled the disagreement by choosing one of the two options selected by the previous coders.

### 3.6. Data collection for physicians’ conflicts of interest

LF obtained the list of the physicians from the CMIT. He then collected the official identifiers of the physicians (RPPS). Using these identifiers, LFhe then downloaded all the fundings for each member of the CMIT from the 1st January of 2013 to the 31st December of 2019 using eurofordocs.fr as in the original study. He then summed these fundings by physician and matched the resulting database with physicians’ attitudes. Fundings from Sanofi include those from Sanofi Pasteur MSD SNC and Sanofi SA.

### 3.7. Data collection for mediators

Researcher’s h-index was collected on Web of Science (webofknowledge.com) by FC. FC also assessed physicians’ affiliation to the IHU Méditerranée-Infection (IHU-MI) by looking, for each physician on the CMIT member list, their current affiliation and whether they appeared on the IHU Méditerrannée-Infection’s website (https://www.mediterranee-infection.com/).

## Appendix 2. Mediation analysis

In the results section of the paper, we found a significant negative correlation between the total amount of money received by physicians from pharmaceutical enterprises and their attitude towards HCQ. One explanation for this result is that, as claimed by Roussel and Raoult (2020), hostility towards HCQ is orchestrated by pharmaceutical enterprises. However, it is not the *only* possible explanation. In this section, we put two alternate explanations to test.

A first alternate explanation is that physicians who demonstrate better research skills might be both (i) more likely to receive fundings from pharmaceutical enterprises, and (ii) more likely to be hostile to HCQ, given the lack of scientific evidence in favor of its efficiency against COVID-19. If so, we should expect physicians’ research skills to mediate the link between their conflict of interest and their attitudes towards HCQ. However, a physician’s research skills are hard to assess. As an imperfect measure (proxy) of physicians’ research skills, we used their h-index, as found on Web of Science. Thus, we hypothesized that the link between physicians’ conflict of interest and their attitude towards HCQ would be mediated by their h-index.^3^

A second alternate explanation, which did not appear in our pre-registration plan, was suggested to us by our exploration of the data. As can be seen in Figure 1a, correlations between physicians’ attitudes towards HCQ and the amount of money they received from Gilead Sciences and pharmaceutical enterprises in general seemed to be mainly driven by a small group of physicians who (i) strongly supported HCQ but (ii) received little money from pharmaceutical enterprises. Wondering what these physicians might have in common, we realized that most of them were members of the IHU Méditerranée-Infection (henceforth: IHU-MI), the research institute directed by Didier Raoult himself.

Thus, one alternate explanation is that the correlation we observed is due to the fact that (i) members of the IHU-MI are more likely to support HCQ, and (ii) members of the IHU-MI are less likely to (directly) receive money from pharmaceutical enterprises. In line with this explanation, we also hypothesized that physicians’ affiliation to IHU-MI (coded as NO = 0, YES = 1) would mediate the link between their conflict of interest and their attitude towards HCQ.

Looking at both potential mediators, we found that physicians’ h-index was neither significantly related with their total conflict of interests (β = 0.16, *p* = 0.26, *R*^*2*^ = 0.03), nor with their attitudes towards HCQ (β = 0.09, *p* = 0.55, *R*^*2*^ < 0.01). As for physicians’ affiliation to the IHU-MI, it significantly predicted their attitude towards HCQ and explained an important part of the variance (β = 0.54, *p* < 0.001, *R*^*2*^ = 0.30). but it did not significantly predict the amount of money received from pharmaceutical enterprises (β = −0.21, *p* = 0.13, *R*^*2*^ = 0.05), though this might be partly due to the fact that our sample of IHU-MI members was small (*N* = 6). This suggests that we might not have enough data to run a proper mediation test.

To investigate whether physicians’ h-index and affiliation to IHU-M mediated the relationship between their conflict of interest and their attitudes towards HCQ, we conducted a multiple regression analysis with attitudes towards HCQ as dependent variable and total conflicts of interests, physicians’ h-index and affiliation to IHU-MI as predictors. Results are presented in Table 5. As can be seen, entering physicians’ h-index and affiliation into the model turned the relationship between total funding and attitudes only marginally significant. However, total funding still explained an interesting part of the variance: a model without total funding (with only h-index and affiliation as predictors) only explains 30% of variance (against 36%) and is marginally worse than the model including total funding from pharmaceutical enterprises (*p* = 0.055).

**Table 4.** Results of multiple regression analysis with physicians’ attitudes towards HCQ (first period) as dependent variable and total conflicts of interests, physicians’ h-index and affiliation to IHU-MI as predictors.

| | B | $\beta$ | SE | t | p |
| --- | --- | --- | --- | --- | --- |
| (Intercept) | 0.34 | - | 0.33 | 1.02 | 0.315 |
| Total funding | $-3.23 \times 10^{-6}$ | -0.25 | $1.68 \times 10^{-6}$ | -1.97 | 0.055 |
| h-index | $-1.57 \times 10^{-3}$ | -0.02 | $9.16 \times 10^{-3}$ | -0.17 | 0.864 |
| Affiliation to IHU-MI | 1.75 | 0.50 | 0.45 | 3.87 | < .001 |
| $R^2 = 0.36$, $F = 8.52$, $p < .001$ | | | | | |

**Table 5.** Results of multiple regression analysis with physicians’ attitudes towards HCQ (first period) as dependent variable and gilead fundings, physicians’ h-index and affiliation to IHU-MI as predictors.

| | b | $\beta$ | SE | t | p |
| --- | --- | --- | --- | --- | --- |
| (Intercept) | 0.31 | - | 0.34 | 0.91 | 0.367 |
| Gilead funding | $-1.02 \times 10^{-5}$ | -0.11 | $1.17 \times 10^{-5}$ | -0.88 | 0.386 |
| h-index | $-4.04 \times 10^{-3}$ | -0.06 | $9.43 \times 10^{-3}$ | -0.43 | 0.670 |
| Affiliation to IHU-MI | 1.88 | 0.53 | 0.47 | 4.04 | < 0.000 |
| $R^2 = 0.31$, $F = 7.03$, $p < .001$ | | | | | |

Given the weight of physicians’ affiliation to IHU-MI in predicting their attitudes towards HCQ, we also investigated the relationship between total conflict of interest and attitudes towards HCQ after excluding members of the IHU-MI from analysis. There was still a marginally significant relationship between the two variables: *r*(42) = −0.29, *p* = 0.054. (X) Thus, even when excluding members of the IHU-MI, total conflict of interests still seemed to (possibly) explain around 9% of variance.

Finally, to investigate whether the marginally significant relationship we observed between conflicts of interests with Gilead Sciences and attitudes towards HCQ could be explained by our two potential mediators, we performed similar analyses with fundings from Gilead as dependent variable. Results from a multiple regression analysis are presented in Table 6.

**Table 6.**
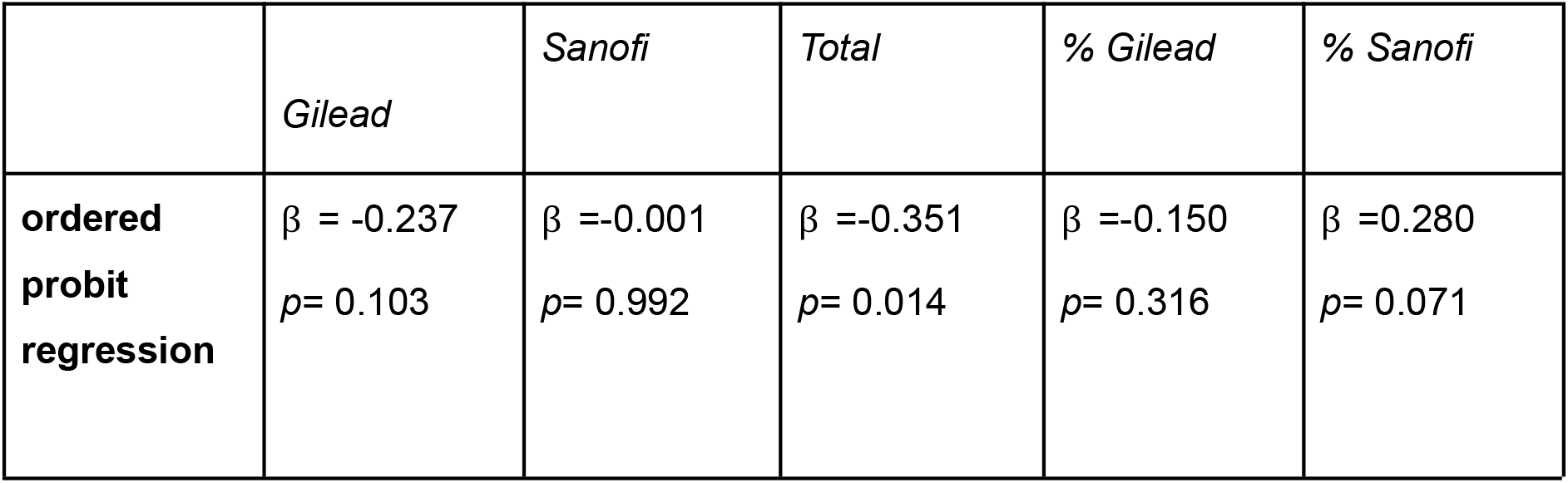
Results of several bivariate ordered probit regressions with physicians’ attitudes towards HCQ (first period) as dependent variable and several measures of fundings as predictors. The β (standardized coefficients) are the fully standardized ones: (bstdXY).

|  | *Gilead* | *Sanofi* | *Total* | *% Gilead* | *% Sanofi* |
| --- | --- | --- | --- | --- | --- |
| **ordered probit regression** | $\beta = -0.237$<br>$p = 0.103$ | $\beta = -0.001$<br>$p = 0.992$ | $\beta = -0.351$<br>$p = 0.014$ | $\beta = -0.150$<br>$p = 0.316$ | $\beta = 0.280$<br>$p = 0.071$ |

The association between gilead funding and attitudes towards HCQ disappears in this setting.

When excluding members of the IHU-MI from analysis, the relationship between these two variables was non-significant (*r*= −0.15, *p* = 0.345), and fundings from Gilead explained only 2% of variance in attitudes.

**Figure S1.**
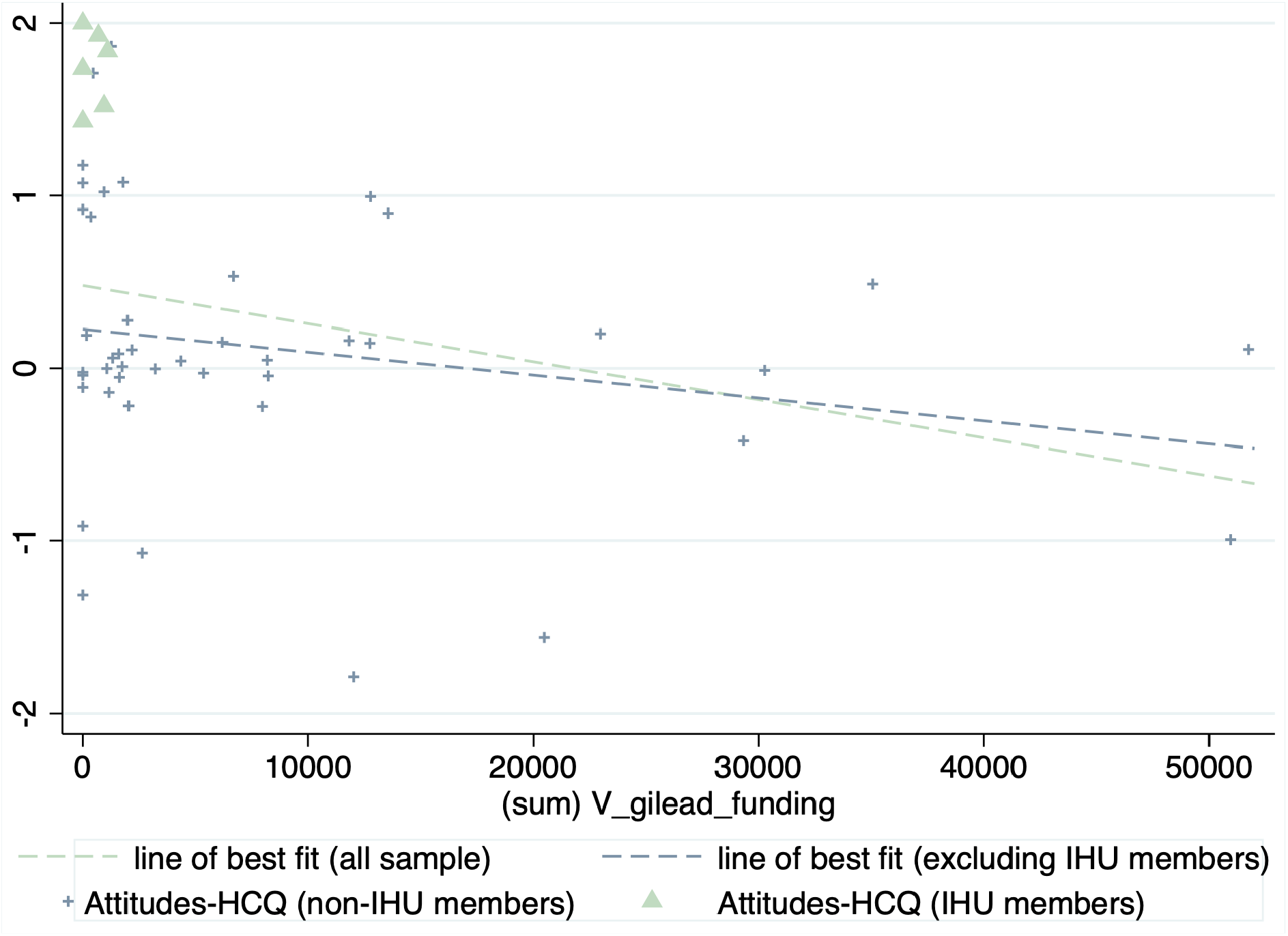
Plotting the relationship between average attitudes towards HCQ and gilead fundings among IHU members and non IHU members. Data was jitterized in order to circumvent overplotting

This figure shows the same pattern as Figure 1 the observed negative relationship between gilead fundings and attitudes is partly driven by IHU members which constitute a group of outliers with both zero fundings and strong expressed support of the molecule.

## APPENDIX 3

We used an ordered probit regression which might be more suited for our discrete outcome. We assessed the relationship between the various measures of fundings and physicians’ attitudes (as in Table 3). The results are presented in the table below.

The conclusions are the same as in our main analysis. While the association between total fundings and opinion is significant in bivariate models, none of the other components of the fundings significantly correlates with opinion. We chose not to display marginal effects, as we are merely interested in the direction of the relationship and its significance.

When excluding the members of the IHU-MI and using ordered probit regressions, the relationship between gilead fundings and attitudes towards HCQ is still non-significant *(p* = 0.412), and that between total fundings and attitudes becomes non or marginally significant (*p* = 0.076) and remains so when controlling for the h-index *(p* = 0.090).

## APPENDIX 4

In Section 5, we conducted correlational analyses using a measure of physicians’ attitudes that relied on the more extreme attitude they expressed, in order to mirror the integer-based score used by Roussel and Raoult (2020). Here, we conducted the same analyses using an average score (i.e. the mean of their different attitudes they expressed) instead. Using these scores, the association between gilead fundings and attitudes towards HCQ becomes significant (*r* = - 0.30, *p* < 0.05). However, our general conclusions remain. There is still a stronger association between total fundings and attitudes towards HCQ (*r* = −0.38, *p* < 0.01,), and the association between the ratio of gilead funding and attitudes towards HCQ is still non-significant (*r* = - 0.15, *p* > 0.1).

**Table 7.** Average physicians’ attitudes towards HCQ (first period) in function of various measures of conflict of interests (two methods: Pearson correlation, Regression). For the %Gilead and %Sanofi column, physicians who received no money from pharmaceutical industries were excluded, leaving us with *N* = 49 physicians. For the %Gilead and %Sanofi column, physicians who received no money from pharmaceutical industries were excluded, leaving us with *N* = 49 physicians.

|  | *Gilead* | *Sanofi* | *Total* | *% Gilead* | *% Sanofi* |
| --- | --- | --- | --- | --- | --- |
| **Pearson** | $r = -0.30$<br>$p = 0.03$ | $r = -0.05$<br>$p = 0.70$ | $r = -0.39$<br>$p = 0.005$ | $r = -0.15$<br>$p = 0.30$ | $r = 0.18$<br>$p = 0.22$ |
| **Regression**<br>(OLS) | $R^2 = 0.09$ | $R^2 = 0.00$ | $R^2 = 0.15$ | $R^2 = 0.02$ | $R^2 = 0.03$ |

## APPENDIX 5

We created three new dummies:

- *pos_opinion* equals 1 if the most extreme opinion is superior to 0 and 0 in the other case on HCQ in period 1 (encompassing both the cases of a different opinion expressed or not having expressed an opinion);
- *neutral_opinion* equals 1 if the physician has expressed a neutral opinion on HCQ in period 1 (the opinion index equals 0);
- *negative_opinion* equals 1 when the physician has expressed a negative opinion on HCQ in period 1(opinion index is inferior to 0).

We then looked at the association between each component of fundings and these indicators using a logit model. This has two merits. Firstly, it allows us to increase our sample size. Secondly, it adds granularity into the analysis by highlighting heterogeneous effects of the fundings across the opinion scale.

**Table 8.** Results of logit models using each of the three dummies as outcomes and physicians’ attitudes towards HCQ as dependent variables. For the %Gilead and %Sanofi column, physicians who received no money from pharmaceutical industries were excluded, leaving us with *N* = 49 physicians. The β (standardized coefficients) are the fully standardized ones: (bstdXY).

|  | *Gilead* | *Sanofi* | *Total* | *% Gilead* | *% Sanofi* |
| --- | --- | --- | --- | --- | --- |
| **positive opinion** | $\beta = -0.110$<br>$p = 0.310$ | $\beta = -0.051$<br>$p = 0.634$ | $\beta = -0.204$<br>$p = 0.085$ | $\beta = 0.083$<br>$p = 0.532$ | $\beta = 0.162$<br>$p = 0.342$ |
| **neutral opinion** | $\beta = -0.092$<br>$p = 0.574$ | $\beta = -0.061$<br>$p = 0.733$ | $\beta = -0.185$<br>$p = 0.381$ | $\beta = -0.044$<br>$p = 0.768$ | $\beta = -0.141$<br>$p = 0.477$ |
| **negative opinion** | $\beta = 0.275$<br>$p = 0.029$ | $\beta = 0.132$<br>$p = 0.261$ | $\beta = -0.444$<br>$p = 0.007$ | $\beta = -0.136$<br>$p = 0.558$ | $\beta = -0.161$<br>$p = 0.582$ |

These results are in line with our core findings. There is an association between Gilead Sciences fundings and negative attitudes towards HCQ, but (i) there is still a much stronger association between attitudes and total fundings, even when excluding IHU-MI members. and (ii) the association between the ratio of gilead funding to total fundings and negative attitudes towards HCQ is still non-significant.

## APPENDIX 6

Because affiliation to IHU seemed to play an important role in the relationship between fundings and attitudes towards HCQ, we used a mixed (multi-level) model to account for the hierarchical structure of the data. When nesting the data at the level of affiliation (members of the IHU-MI or not), the association between gilead fundings and average attitudes towards HCQ is marginally significant (b=-1.4*10^-5, p=0.096). The association between total fundings and average attitudes is significant (b= −3.07*10^-6, p=0.011).

With the same hierarchical model, the association between gilead fundings and their more extreme attitudes towards HCQ is still non-significant (b=-1.25*10^-5, p=0.260). The association between total fundings and attitudes remains significant as well when considering the most extreme attitudes (b= −3.56*10^-6, p=0.026).

## APPENDIX 7

The aim of this appendix is to check whether h_index and gilead fundings could be suitable mediators of the significant relationship we found in Annex 1 between gilead fundings and average attitudes towards HCQ. It is not the case. Neither of the two variables are correlated both with attitudes and gilead fundings.

As in the core of the article, we computed the association in the subsamples of physicians who expressed an opinion.

**Table 9.** Testing whether h_index could be a suitable mediator to gilead funding.

| Associations | Regression results (OLS) |
| --- | --- |
| **gilead_funding - h_index**<br>*(excluding members who did not express an opinion)* | $\beta=0.152$<br>$p= 0.292$ |
| **average attitude toward HCQ in period 1**<br>**toward - h_index** | $\beta=0.059$<br>$p=0.683$ |

**Table 10.**
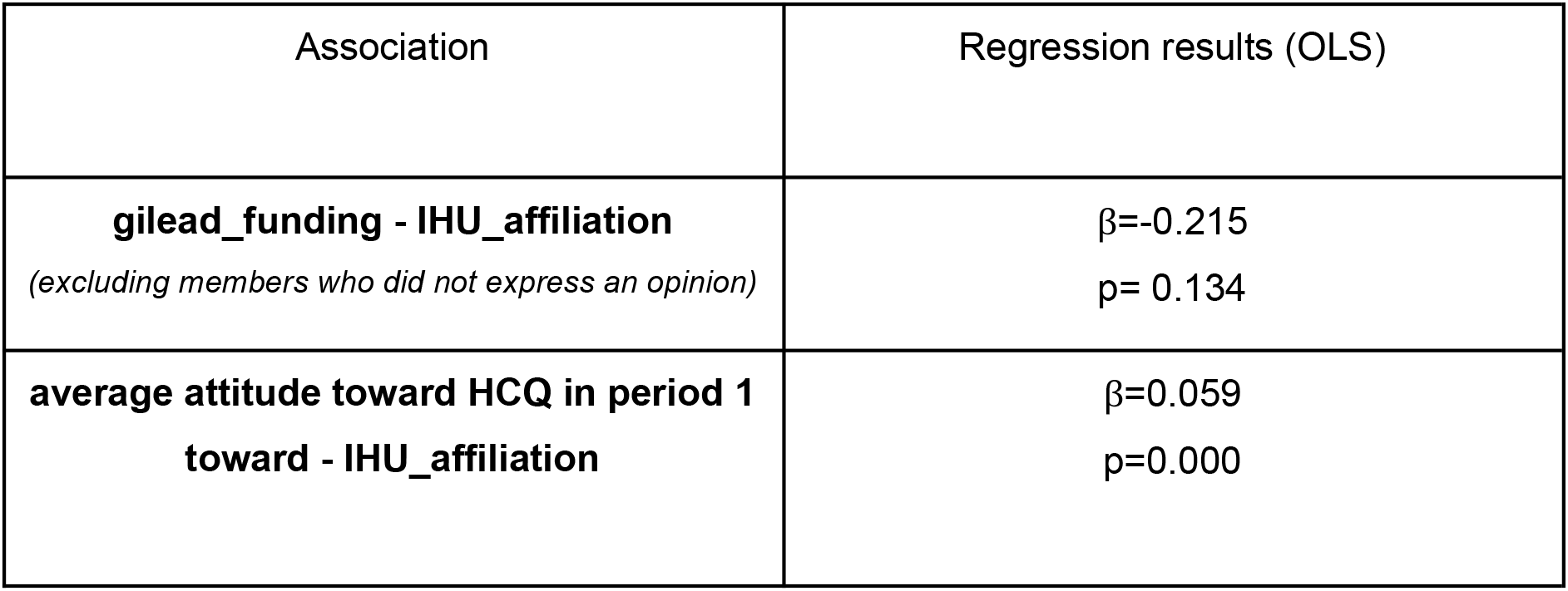
Testing whether IHU_affiliation could be a suitable mediator to gilead funding.

| Association | Regression results (OLS) |
| --- | --- |
| **gilead_funding - IHU_affiliation**<br>*(excluding members who did not express an opinion)* | $\beta=-0.215$<br>$p= 0.134$ |
| **average attitude toward HCQ in period 1**<br>**toward - IHU_affiliation** | $\beta=0.059$<br>$p=0.000$ |

## APPENDIX 8

Consistently with our pre-registration plan, we also tested whether physicians who received more money from the pharmaceutical industry or from specific pharmaceutical companies were most likely to express an attitude towards HCQ.

To remain consistent with our previous analyses, we focused on one outcome: a dummy which equals 1 if the physician has expressed an opinion about HCQ in the first period. The results are clear-cut: there does not seem to be any type of association between fundings and propensity to express one’s attitudes towards HCQ.

**Table 11.** Relationship between different measures of fundings and whether physicians expressed an attitude about HCQ, estimated through three different methods (logit model, probit model, and linear probability model).

|  | *Gilead* | *Sanofi* | *Total* | *% Gilead* | *% Sanofi* |
| --- | --- | --- | --- | --- | --- |
| **Logit model** | $\beta = 0.121$<br>$p = 0.253$ | $\beta = 0.203$<br>$p = 0.208$ | $\beta = 0.128$<br>$p = 0.262$ | $\beta = -0.122$<br>$p = 0.272$ | $\beta = -0.031$<br>$p = 0.772$ |
| **Probit model** | $\beta = 0.137$<br>$p = 0.250$ | $\beta = 0.231$<br>$p = 0.186$ | $\beta = 0.145$<br>$p = 0.251$ | $\beta = -0.134$<br>$p = 0.267$ | $\beta = -0.036$<br>$p = 0.770$ |
| **Linear Probability Model** | $\beta = 0.109$<br>$p = 0.247$ | $\beta = 0.141$<br>$p = 0.131$ | $\beta = -0.110$<br>$p = 0.244$ | $\beta = -0.105$<br>$p = 0.270$ | $\beta = -0.027$<br>$p = 0.774$ |

## APPENDIX 9

Consistently with our pre-registration plan, we also included three other variables in our dataset: age (which was simply estimated by computing 2021 - their year of birth), whether the physician is a professor or a mere medical doctor, and the number of publications registered on *Web of Science (WoS)*. Our pre-registration plan mentioned publications registered on pubmed. However, there are no author profiles on *pubmed*. Thus, we turned to WoS which tracks and counts the number of publications by physicians. While we also registered the possibility to lead the analyses again on the subsample of physicians with scientific publications, all the infectiologists on the list authored or co-authored at least one publication. Hence, it is impossible to split the sample into non-researchers and researchers without setting an arbitrary threshold of scientific articles.

### Appendix 9.1 Controlling for these variables does not change our results

Adding these variables as controls in multiple linear regressions does not change our conclusions. The association between gilead fundings and average opinion disappears when controlling for other variables, especially IHU_affiliation. The association between total fundings and public expression of physicians resists pretty well to the addition of all these controls.

**Table 12.**
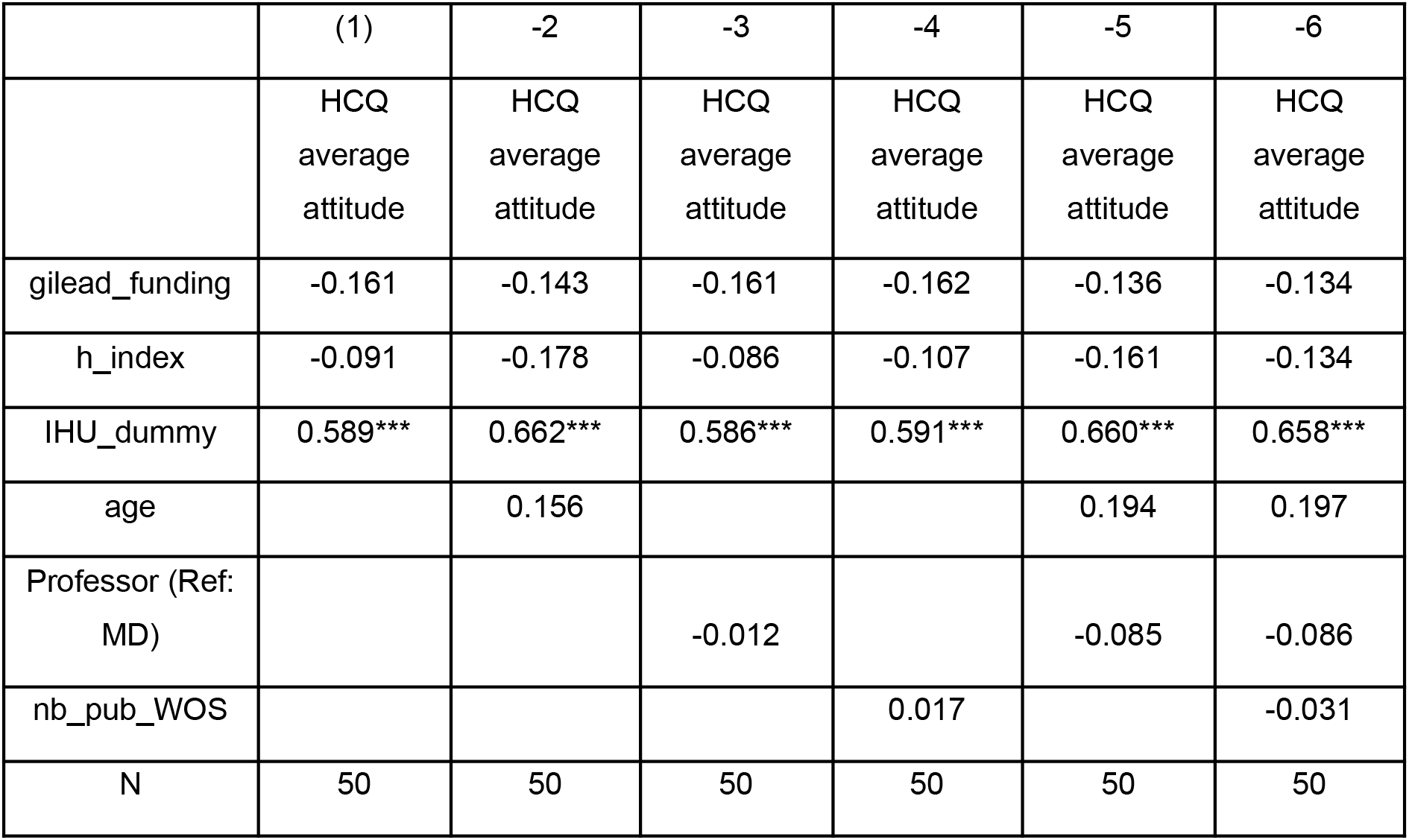
Coefficients are beta standardized. * p<0.10, ** p<0.05, *** p<0.01

**Table 13.** Coefficients are beta standardized. * p<0.10, ** p<0.05, *** p<0.01

|  | -1 | -2 | -3 | -4 | -5 | -6 |
| --- | --- | --- | --- | --- | --- | --- |
|  | HCQ<br>average<br>attitude | HCQ<br>average<br>attitude | HCQ<br>average<br>attitude | HCQ<br>average<br>attitude | HCQ<br>average<br>attitude | HCQ<br>average<br>attitude |
| V_total_fu~g | -0.257** | -0.255** | -0.257** | -0.289** | -0.253** | -0.276** |
| h_index | -0.065 | -0.158 | -0.059 | -0.219 | -0.137 | -0.243 |
| IHU_dummy | 0.561*** | 0.638*** | 0.558*** | 0.572*** | 0.632*** | 0.636*** |
| age |  | 0.174 |  |  | 0.215 | 0.204 |
| Pr |  |  | -0.014 |  | -0.093 | -0.088 |
| nb_pub_WOS |  |  |  | 0.177 |  | 0.127 |
| N | 50 | 50 | 50 | 50 | 50 | 50 |

### Appendix 9.2 testing whether these two news variables could be suitable mediators for gilead fundings and total fundings

None variables are suitable mediators for the respective relationship between the two fundings components and the average opinion on HCQ.

Age is not correlated with total fundings (when excluding physicians who did not express an opinion) (*p*=0.242) nor with the average opinion towards HCQ (*p*=0.446). Age is also not correlated with gilead fundings (*p*=0.622) - when excluding physicians who did not express an opinion.

Whether the physician is a professor or a doctor is not correlated with total fundings (with the same necessary sample restriction) (*p*=0.326) nor with the average opinion towards HCQ (*p*=0.336). This dummy variable is also not correlated with gilead fundings (*p*=0.290) - again, we exclude physicians who did not give their opinion on HCQ.

Finally, the number of publications registered on Web of Science is correlated to the fundings in the subsample of physicians who gave their opinion (*p*=0.019). However, this variable is not associated with the average opinion on HCQ (*p*=0.894). The number of registered publications on WOS is not correlated with gilead fundings (0.116) - again, the analysis includes only the physicians who gave their opinion.

## Footnotes

1 In media, the terms “chloroquine” and “hydroxychloroquine” tended to be used interchangeably. Thus, when it was clear that “chloroquine” was used to refer to “hydroxychloroquine” or the protocol recommended by Didier Raoult, we coded these interventions as relevant.

2 For the ‘very favourable’ and ‘very unfavourable’ categories, we forgot to include the passage about “claiming that the treatment works/doesn’t work against COVID-19” in the preregistration. However, we think it is quite obvious that such interventions should fall into these categories.

3 In our pre-registration, we also mentioned age and universitary status as potential mediators. We run these mediation analyses in appendix 9.

## Notes

### Competing Interest Statement

The authors have declared no competing interest.

### Funding Statement

All the authors but one did not receive fundings for this project. Florian Cova received a grant from the Swiss National Science Foundation: "Eudaimonic emotions and the (meta)philosophy of well-being"

### Author Declarations

The study includes funding data of physicians which is openly available on eurofordocs.eu. The list of physicians is available here: https://www.infectiologie.com/fr/organigramme-cmit.html

### Summary of Updates

corrected a typo

## References

[1] Fuhrer, J., & Cova, F. (2020). “Quick and dirty”: Intuitive cognitive style predicts trust in Didier Raoult and his hydroxychloroquine-based treatment against COVID-19. Judgment & Decision Making, 15(6), 889–908.

[2] Gould, S., & Norris, S.L. (2021). Contested effects and chaotic policies: the 2020 story of (hydroxy) chloroquine for treating COVID-19. Cochrane Database of Systematic Review, 2021(3), ED000151.

[3] Schultz, É., & Ward, J. K. (in press). Science under Covid-19’s magnifying glass: Lessons from the first months of the chloroquine debate in the French press. Journal of Sociology.

[4] Rosendaal, F. R. (2020). Review of:“Hydroxychloroquine and azithromycin as a treatment of COVID-19: results of an open-label non-randomized clinical trial Gautret et al 2010. International Journal of Antimicrobial Agents, 56(1), 106063.

[5] Fiolet, T., Guihur, A., Rebeaud, M. E., Mulot, M., Peiffer-Smadja, N., & Mahamat-Saleh, Y. (2021). Effect of hydroxychloroquine with or without azithromycin on the mortality of coronavirus disease 2019 (COVID-19) patients: a systematic review and meta-analysis. Clinical Microbiology and Infection, 27(1), 19–27.

[6] Singh, B., Ryan, H., Kredo, T., Chaplin, M., & Fletcher, T. (2021). Chloroquine or hydroxychloroquine for prevention and treatment of COVID-19. Cochrane Database of Systematic Reviews, 2021(2), CD013587.

[7] Roussel, Y., Gautret, P., & Raoult, D. (2021). The War against chloroquine. Unpublished manuscript, IHU Méditerranée-Infection. Last retrieved at: https://www.mediterranee-infection.com/wp-content/uploads/2020/04/Guerre-chloroquine.pdf

[8] Chabrière, E. (2021). Does IHU-Méditerranée Infection influence Gilead Sciences’ stock price? New Microbes and New Infections, 39, 100711.

[9] Roussel, Y., & Raoult, D. (2020). Influence of conflicts of interest on public positions in the COVID-19 era, the case of Gilead Sciences. New Microbes and New Infections, 38, 100710.

[10] Piepho, H-P. “‘Influence of conflicts of interest on public positions in the COVID-19 era, the case of Gilead Sciences’ by Roussel and Raoult (2020).” New Microbes and New Infections, 37, 100730.

[11] Raoult, D. (2020). La science est un sport de combat. Humensciences

